# Estimating treatment effect for individuals with progressive multiple sclerosis using deep learning

**DOI:** 10.1101/2021.10.31.21265690

**Authors:** Jean-Pierre R. Falet, Joshua Durso-Finley, Brennan Nichyporuk, Julien Schroeter, Francesca Bovis, Maria-Pia Sormani, Doina Precup, Tal Arbel, Douglas Lorne Arnold

## Abstract

Progressive forms of multiple sclerosis (MS) remain resistant to treatment. Since there are currently no suitable biomarkers to allow for phase 2 trials, pharmaceutical companies must proceed directly to financially risky phase 3 trials, presenting a high barrier to drug development. We address this problem through predictive enrichment, which randomizes individuals predicted to be most responsive in order to increase a study’s power. Specifically, deep learning is used to estimate conditional average treatment effect (CATE) using baseline clinical and imaging features, and rank individuals on the basis of their predicted response to anti-CD20 antibodies. We leverage a large dataset from six different randomized clinical trials (n = 3, 830). In a left-out test set of primary progressive patients from two anti-CD20-antibodies trials, the average treatment effect was significantly greater for the 50% (HR, 0.492; 95% CI, 0.266-0.912; *p* = 0.0218) and the 30% (HR, 0.361; 95% CI, 0.165-0.79; *p* = 0.008) predicted to be most responsive, compared to 0.743 (95% CI, 0.482-1.15; *p* = 0.179) for the entire group. The same model could also identify responders to laquinimod, which has a different mechanism of action. We demonstrate important increases in power that would result from the use of this model for predictive enrichment, enabling short proof-of-concept trials.

## 1 Introduction

Several disease modifying therapies have been developed for the treatment of the inflammatory manifestations of relapsing-remitting multiple sclerosis (RRMS) (clinical relapses and lesion activity) using the strategy of performing relatively short and small phase 2 trials with a magnetic resonance imaging (MRI) endpoint for establishing proof of concept and finding the optimal dose, before proceeding to longer, more expensive phase 3 trials. The absence of analogous MRI endpoints for progressive multiple sclerosis has hampered progress in developing drugs for this clinical phase of the disease. While brain atrophy has frequently been used as a surrogate outcome measure in phase 2 trials, there has yet to be an example of a follow-up phase 3 clinical trial that confirms an effect detected in a phase 2 trial where brain atrophy was the primary outcome. As proceeding directly to large, phase 3 trials is expensive and risky, most programs that followed this path have failed to adequately demonstrate efficacy.

It is often the case that medications are more effective in some patients than others. Selecting such a subgroup for inclusion in a clinical trial in order to increase its power is a technique called *predictive enrichment* [1]. A drug proven to be efficacious in a trial enriched with predicted responders can later be tested more confidently in a population predicted to be less responsive. This sequence prevents efficacious medications from having their effect diluted in early clinical trials due to inclusion of a population that is too heterogeneous, while still allowing for broadening of indication criteria. It also improves the balance of risks and benefits for participants, since those who are unlikely to benefit from a drug would not be exposed to it and therefore would not experience potential adverse effects. A relevant application of predictive enrichment was described by Bovis *et al*. [2], who used Cox Proportional Hazards Cox proportional hazards (CPH) models to successfully predict a more responsive sub-group of RRMS patients to laquinimod, a medication whose average treatment effect in the original phase 3 studies was insufficient for drug approval.

Deep learning is a highly expressive and flexible type of machine learning that can potentially uncover complex, non-linear relationships between baseline patient characteristics and their responsiveness to treatment. However, contrary to traditional machine learning problems where a mapping between features and targets is learned from a sample of observations, the target in a treatment response (or treatment effect) task is not directly observable. Adaptations to machine learning frameworks must therefore be made in order to frame the problem through the lens of causal inference (reviewed in detail in the survey on uplift modeling by Gutierrez & Gérardy [3]). Arguably some of the most popular methods have been tree-based approaches (see Radcliffe & Surry [4] for an example) which model treatment effect directly, and meta-learning approaches [5] which decompose the treatment effect estimation problem into simpler problems that can be tackled using traditional machine learning models. In a recent paper, our group presented a meta-learning approach for the estimation of treatment effect (as measured by suppression of new/enlarging T2-lesions) in RRMS using baseline brain MRI and clinical variables [6].

In this work, we present a new deep learning framework to estimate an individual’s treatment effect using readily available clinical information (demographic characteristics and clinical disability scores) and scalar MRI metrics (lesional and volumetric) obtained at the screening visit of a clinical trial. This approach, based on an ensemble of multi-headed multilayer perceptrons (MLPs), can identify more responsive individuals to both anti-CD20 monoclonal antibodies (anti-CD20-Abs) and laquinimod better than alternative strategies. We demonstrate how using this model for predictive enrichment could greatly improve the feasibility of short proof-of-concept trials in primary progressive multiple sclerosis (PPMS), accelerating therapeutic advances.

## 2 Results

### 2.1 Datasets

Data were pooled from six randomized clinical trials (*n* = 3, 830): OPERA I [7], OPERA II [7], BRAVO [8], ORATORIO [9], OLYMPUS [10], and ARPEGGIO [11] (ClinicalTrials.gov numbers, NCT01247324, NCT01412333, NCT00605215, NCT01194570, NCT00087529, NCT02284568, respectively). OPERA I/II, and BRAVO were RRMS trials which compared ocrelizumab with subcutaneous interferon beta-1a (IFNb-1a), and laquinimod with both intramuscular IFNb-1a and placebo, respectively. ORATORIO, OLYMPUS, and ARPEGGIO were placebo-controlled PPMS trials which studied ocrelizumab, rituximab, and laquinimod, respectively.

The dataset is divided into three subsets for different phases of training and evaluation. The first subset (*n* = 2, 520) contains data from the three RRMS trials, and is used for pre-training the MLP to learn predictors of treatment effect under the RRMS condition (for details, see Section 5, Online Methods). This pre-training phase falls under the umbrella of *transfer learning*, a deep learning strategy that is used to transfer knowledge acquired from a related task to a task with fewer samples in order to improve learning on the latter [12]. The second subset consists of two PPMS trials (*n* = 992): OLYMPUS and ORATORIO. This subset is divided into a 70% training set (*n* = 695) which is used to fine-tune the pre-trained MLP to estimate treatment effect to anti-CD20-Abs, and the remaining 30% (*n* = 297) is left out as a test set to estimate the generalization error of the fully trained model. The third subset contains PPMS data from the trial ARPEGGIO (*n* = 318), which is also left out as a second test set.

Mean and standard deviation for the baseline features and the outcome metrics in the PPMS subset are shown in Table 1, separated by treatment arm (the same statistics for the RRMS subset are shown in Supplementary Table 1). The groups are comparable for all features except for disease duration which is shorter in ORATORIO, and Gad count and T2 lesion volume, which are greater in ORATORIO. This may be due to ORATORIO’s inclusion criteria, which had a maximum time from symptom onset, and to inter-trial differences in automatic lesion segmentation, which are accounting for using a scaling procedure explained in Section 5.1. Some heterogeneity exists between the outcomes of each trial when looking at the placebo arms, which on average have a smaller restricted mean survival time (RMST) at 2 years in ARPEGGIO and OLYMPUS compared to ORATORIO, indicating more rapid disability progression on the Expanded Disability Status Scale (EDSS).

**Table 1:**
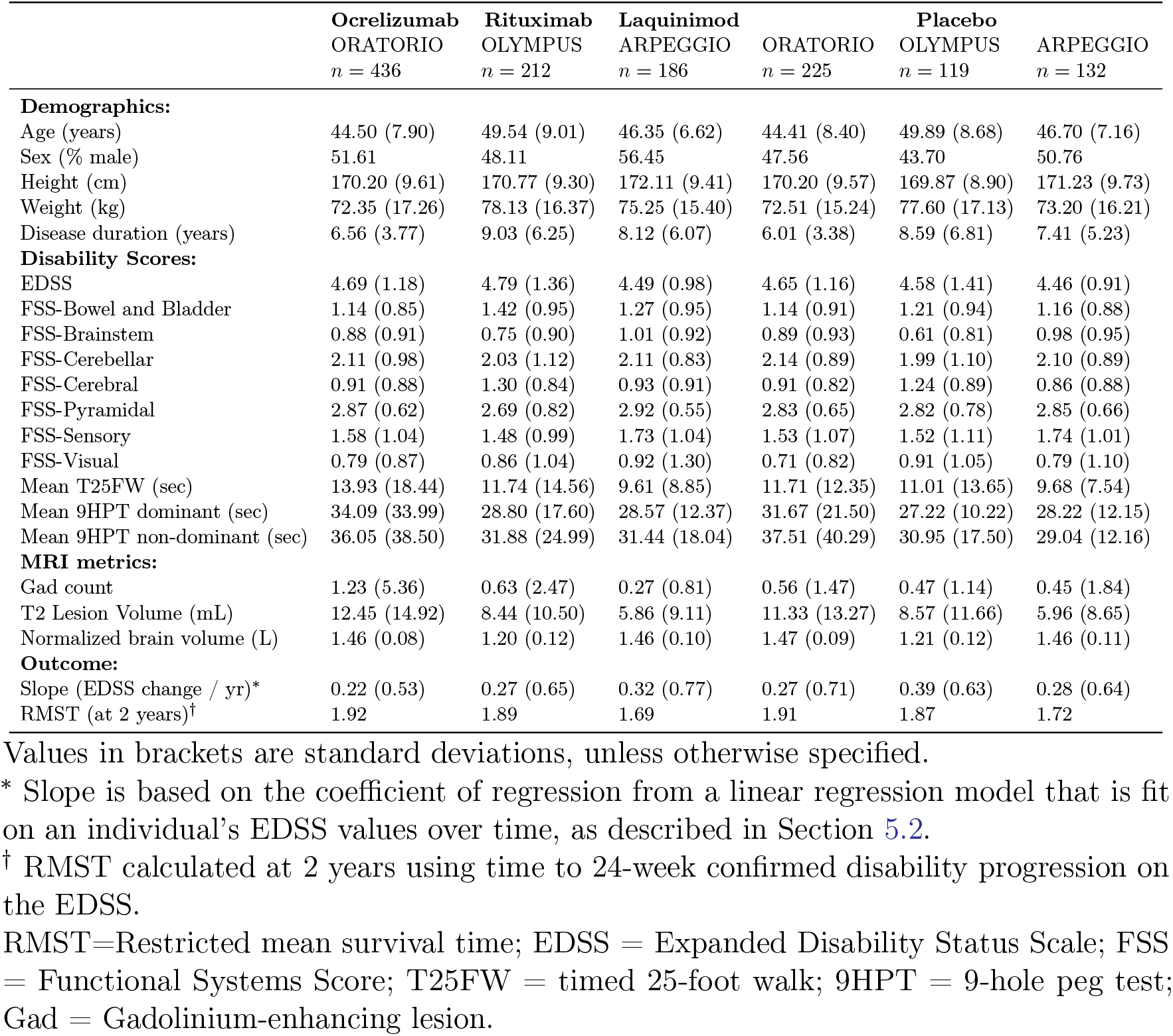
Baseline features and outcomes per treatment arm.

### 2.2 Predicting response to anti-CD20 monoclonal antibodies

As described in Section 5 (Online Methods), we train an ensemble of multi-headed MLPs to predict the change in EDSS over time (obtained by fitting a linear regression model to an individual’s EDSS values recorded over time and taking the slope of the regression to be the prediction target) on both anti-CD20-Abs and placebo. These two predictions are then subtracted to obtained an estimate of the conditional average treatment effect (CATE) for each individual, given their baseline features. A histogram of predictions on the unseen anti-CD20-Abs test set (30% of the dataset, *n* = 297) from the fully trained model is shown in Supplementary Fig. 1. The model’s ability to rank response is assessed using an average difference curve, *AD*(*c*), which is described by Zhao *et al*. [13] and is well suited for measuring performance in predictive enrichment. Our implementation measures the ground-truth average difference in RMST (calculated at 2 years from time to 24-week confirmed disability progression (CDP24)) between anti-CD20-Abs and placebo for individuals predicted to respond more than a certain threshold, as a function of this threshold. The *AD*(*c*) curve for our model, shown in Fig. 1, appropriately increases as a sub-group that is predicted to be more and more responsive is selected. The *AD*_*wabc*_, a metric derived from the area under the *AD*(*c*) curve in Supplementary Methods 3, provides a measure of how well the model can rank individuals on the basis of their responsiveness to treatment. Larger positive *AD*_*wabc*_ values indicate better performance. The *AD*_*wabc*_ in this case is positive and relatively large (0.0565) and nearly monotonic (Spearman r correlation coefficient 0.943), demonstrating the ability for the model to rank response to anti-CD20-Abs.

**Figure 1:**
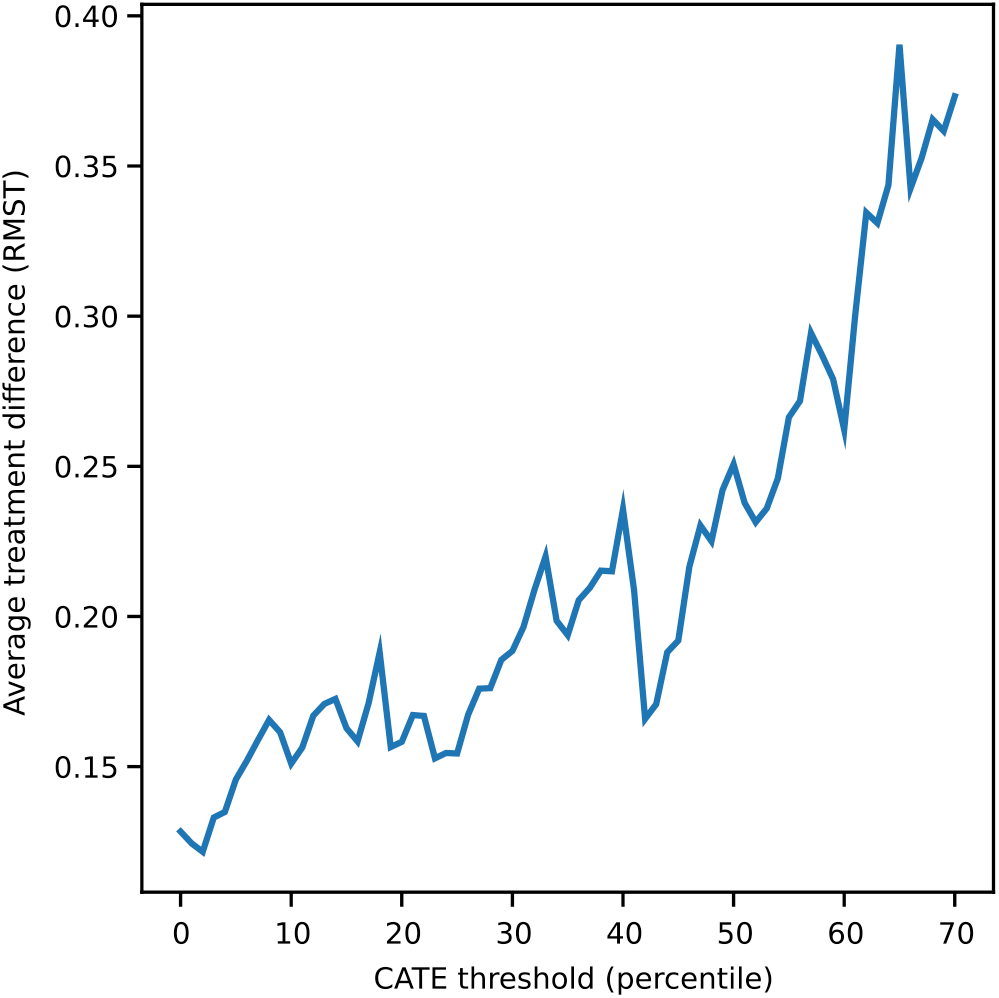
Average treatment difference curve for the anti-CD20-Abs test set. Represents the difference in the ground-truth restricted mean survival time (RMST), calculated at 2 years using time-to-CDP24, between anti-CD20-Abs and placebo, among predicted responders defined using various thresholds. The CATE percentile threshold is the minimum CATE (expressed as a percentile among all CATE estimates in the test set) that is used to define an individual as a responder (i.e. a threshold of 0.7 means the 30% predicted to be most responsive are considered responders).

Kaplan-Meyer curves of the ground-truth time-to-CDP24 in predicted responders and non-responders are shown in Fig. 2 for two predictive enrichment levels (selecting the 50% and the 30% predicted to be most responsive), along with their complement (the 50% and 70% predicted to be least responsive). Compared to the entire test set whose HR is 0.743 (95% CI, 0.482-1.15; *p* = 0.179), predictive enrichment leads to a HR of 0.492 (95% CI, 0.266-0.912; *p* = 0.0218) and 0.361 (95% CI, 0.165-0.79; *p* = 0.008) when selecting the 50% and 30% most responsive, respectively. The corresponding non-responder groups have a HR of 1.11 (95% CI, 0.599-2.05; *p* = 0.744) and 0.976 (95% CI, 0.578-1.65; *p* = 0.925) when selecting the 50% and 70% least responsive, respectively. This heterogeneity suggests that a significant part of the trend for an effect at the whole-group level may be explained by a small proportion of more responsive patients.

**Figure 2:**
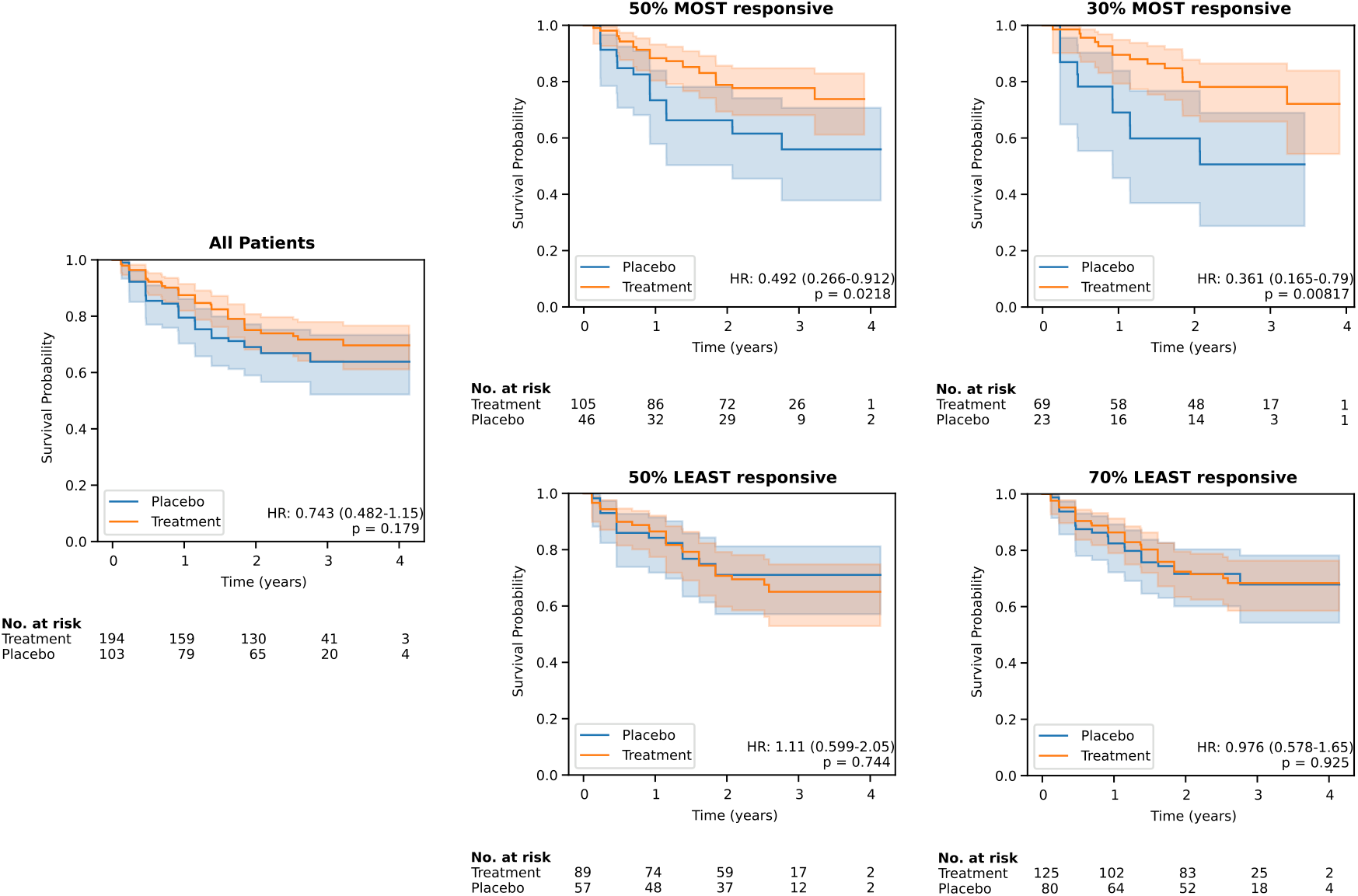
Kaplan-Meyer curves for predicted responders and non-responders to anti-CD20-Abs, defined at two thresholds of predicted effect size. These are compared to the whole group (left). Survival probability is measured in terms of time-to-CDP24 using the EDSS. *p* values are calculated using log-rank tests. Kaplan-Meyer curve 95% confidence intervals are estimated using Greenwood’s Exponential formula.

Of ocrelizumab and rituximab, only the former had a significant effect in a phase 3 trial (ORATORIO), and it is the only drug approved in PPMS. We therefore verified whether the model’s enrichment capabilities are maintained within the ORATORIO subgroup (*n* = 188) of the test set, which has HR of 0.661 (95% CI 0.383-1.14, *p* = 0.135). If selecting the 50% and 30% predicted to be most responsive, the HR reduces to 0.516 (95% CI, 0.241-1.1; *p* = 0.084) and 0.282 (95% CI, 0.105-0.762; *p* = 0.0082), respectively. The corresponding 50% and 70% predicted to be least responsive have a HR of 0.849 (95% CI, 0.385-1.87; *p* = 0.685) and 0.915 (95% CI, 0.471-1.78; *p* = 0.791), respectively.

We then considered specific demographic subgroups to understand their effect on model performance. For men, the model achieved a *AD*_*wabc*_ of 0.0405, while for women the model performs better (*AD*_*wabc*_ = 0.0844). For those with an age < 51, the *AD*_*wabc*_ of 0.0353 is lower than for those with an age >= 51 (*AD*_*wabc*_ = 0.0661). For those with a disease duration < 5, the model performs less well than on those with a disease duration >=5 (*AD*_*wabc*_ = 0.0385 compared to 0.0117). Finally, the model performs better for those with an EDSS < 4.5 (*AD*_*wabc*_ = 0.069) than for those with an EDSS of >= 4.5 (*AD*_*wabc*_ = 0.0451).

Group characteristics for the predicted responders and non-responders, defined at the 50th and 70th percentile thresholds, are shown in Table 2. We observe enrichment across a broad range of input features in the responder sub-groups: younger age, shorter disease duration, higher disability scores, and more lesional activity (particularly T2 lesion volume). The largest effect on the Functional Systems Scores (FSS) was seen in Cerebellar and Visual sub-scores, while FSS-Bowel and Bladder, Brainstem, Cerebral, Pyramidal, and Sensory did not reach statistical significance (*p <* 0.05). Timed 25-foot walk (T25FW) was significantly different only for the 70th percentile threshold. Normalized brain volume was the only baseline MRI feature which did not differ significantly between the two groups at either threshold.

**Table 2:** Group statistics for predicted responders and non-responders to anti-CD20-Abs at the 50th and 70th percentile thresholds.

|  | 50th percentile threshold* |  |  |  | 70th percentile threshold* |  |  |  |
| --- | --- | --- | --- | --- | --- | --- | --- | --- |
|  | Responders | Non-responders | Effect size (95% CI)<sup>†</sup> | *p* value<sup>‡</sup> | Responders | Non-responders | Effect size (95% CI)<sup>†</sup> | *p* value<sup>‡</sup> |
| **Trial contribution:** |  |  |  |  |  |  |  |  |
| OLYMPUS | 55 | 54 |  |  | 35 | 74 |  |  |
| ORATORIO | 96 | 92 |  |  | 57 | 131 |  |  |
| **Demographics:** |  |  |  |  |  |  |  |  |
| Age (years) | 45.20 (8.58) | 47.84 (7.89) | -2.64 (-4.53, -0.76) | 0.006 | 44.59 (9.05) | 47.36 (7.87) | -2.77 (-4.93, -0.61) | 0.013 |
| Sex (% male) | 47.02 | 50.68 | 0.86 (0.53, 1.40) | 0.562 | 45.65 | 50.24 | 0.83 (0.49, 1.40) | 0.530 |
| Height (cm) | 170.05 (10.56) | 170.55 (8.80) | -0.50 (-2.72, 1.71) | 0.657 | 169.78 (10.29) | 170.52 (9.47) | -0.74 (-3.23, 1.75) | 0.560 |
| Weight (kg) | 76.17 (18.93) | 72.96 (13.77) | 3.21 (-0.56, 6.98) | 0.096 | 75.68 (20.07) | 74.10 (14.87) | 1.58 (-3.04, 6.20) | 0.502 |
| Disease duration (years) | 6.07 (4.14) | 8.72 (5.45) | -2.65 (-3.76, -1.54) | <0.001 | 5.79 (4.15) | 8.09 (5.19) | -2.30 (-3.41, -1.19) | <0.001 |
| **Disability Scores:** |  |  |  |  |  |  |  |  |
| EDSS | 4.87 (1.18) | 4.52 (1.23) | 0.34 (0.07, 0.62) | 0.015 | 5.07 (1.14) | 4.53 (1.21) | 0.54 (0.25, 0.83) | <0.001 |
| FSS-Bowel and Bladder | 1.25 (0.93) | 1.11 (0.80) | 0.14 (-0.05, 0.34) | 0.157 | 1.27 (0.98) | 1.15 (0.82) | 0.12 (-0.11, 0.35) | 0.315 |
| FSS-Brainstem | 0.82 (0.93) | 0.79 (0.87) | 0.04 (-0.17, 0.24) | 0.726 | 0.90 (0.95) | 0.77 (0.88) | 0.13 (-0.10, 0.36) | 0.265 |
| FSS-Cerebellar | 2.38 (0.97) | 1.78 (1.05) | 0.60 (0.37, 0.83) | <0.001 | 2.57 (0.81) | 1.86 (1.08) | 0.71 (0.48, 0.93) | <0.001 |
| FSS-Cerebral | 1.07 (0.83) | 1.05 (0.89) | 0.02 (-0.18, 0.22) | 0.848 | 1.13 (0.84) | 1.04 (0.87) | 0.09 (-0.12, 0.30) | 0.404 |
| FSS-Pyramidal | 2.75 (0.69) | 2.90 (0.58) | -0.14 (-0.29, 0.00) | 0.052 | 2.77 (0.76) | 2.85 (0.58) | -0.08 (-0.26, 0.10) | 0.382 |
| FSS-Sensory | 1.55 (1.06) | 1.64 (1.02) | -0.08 (-0.32, 0.15) | 0.488 | 1.56 (1.00) | 1.61 (1.06) | -0.05 (-0.30, 0.20) | 0.703 |
| FSS-Visual | 1.04 (1.04) | 0.43 (0.62) | 0.62 (0.42, 0.81) | <0.001 | 1.28 (1.07) | 0.50 (0.71) | 0.78 (0.54, 1.02) | <0.001 |
| Mean T25FW (sec) | 13.55 (17.61) | 10.75 (11.08) | 2.80 (-0.55, 6.15) | 0.103 | 15.95 (21.79) | 10.48 (9.82) | 5.47 (0.77, 10.17) | 0.024 |
| Mean 9HPT dominant (sec) | 32.62 (26.89) | 26.70 (10.24) | 5.92 (1.29, 10.55) | 0.013 | 36.01 (33.25) | 26.88 (9.89) | 9.13 (2.12, 16.15) | 0.012 |
| Mean 9HPT non-dominant (sec) | 37.33 (31.11) | 26.97 (9.32) | 10.36 (5.14, 15.58) | <0.001 | 42.39 (38.33) | 27.68 (9.33) | 14.71 (6.68, 22.75) | <0.001 |
| **MRI metrics:** |  |  |  |  |  |  |  |  |
| Gad count | 1.62 (3.14) | 0.16 (0.48) | 1.46 (0.95, 1.97) | <0.001 | 1.90 (3.64) | 0.46 (1.27) | 1.44 (0.67, 2.22) | <0.001 |
| T2 Lesion Volume (mL) | 13.09 (12.85) | 7.72 (10.17) | 5.37 (2.73, 8.01) | <0.001 | 14.31 (14.22) | 8.72 (10.27) | 5.59 (2.33, 8.85) | <0.001 |
| Normalized brain volume (L) | 1.37 (0.16) | 1.38 (0.16) | -0.02 (-0.05, 0.02) | 0.367 | 1.35 (0.16) | 1.38 (0.16) | -0.03 (-0.07, 0.01) | 0.107 |

Values in brackets are standard deviations, unless otherwise specified.
\*Percentile threshold for defining responders. The 50th percentile defines responders as the top 50% who are predicted to be most responsive, while the 70th percentile defines them as the top 30%. The non-responders are those who fall below the percentile threshold.
<sup>†</sup>Effect size is the average difference between responders and non-responders for all covariates except for “sex” which is an odd’s ratio (OR).
<sup>‡</sup>*p* values for continuous and ordinal variables are calculated using a two-sided Welch’s t-test due to unequal variances/sample sizes. *p* value for the categorical variable “sex” is calculated using a two-sided Fisher’s exact test due to unequal and relatively small sample sizes.
EDSS = Expanded Disability Status Scale; FSS = Functional Systems Score; T25FW = timed 25-foot walk; 9HPT = 9-hole peg test; Gad = Gadolinium-enhancing lesion.

### 2.3 Predicting response to laquinimod

To determine whether the same model trained on the anti-CD20-Abs dataset could be predictive of treatment response to a medication with a different mechanism of action, and to provide a second validation for the model trained on the single 70% training set in the first anti-CD20-Abs experiment, we tested it on data from ARPEGGIO (*n* = 318). The model trained on the anti-CD20-Abs training dataset also generalized to this second unseen test set, as shown by a positive *AD*_*wabc*_ = 0.0208. From the whole-group HR of 0.667 (95% CI: 0.369-1.2; *p* = 0.933), selecting the 50% and the 30% predicted to be most responsive yields a HR of 0.492 (95% CI 0.219-1.11; *p* = 0.0803) and 0.338 (95% CI, 0.131-0.872; *p* = 0.0186) for the top 50% and 30% predicted to be most responsive, respectively. The corresponding 50% and 70% predicted to be least responsive have a HR of 0.945 (95% CI, 0.392-2.28; *p* = 0.901) and 0.967 (95%CI, 0.447-2.09; *p* = 0.933), respectively. The Kaplan-Meyer curves for these predicted subgroups are shown in Supplementary Fig. 2.

Group characteristics for predicted responders are shown in Supplementary Table 2. Groupwise differences are largely similar to those obtained on the anti-CD20-Abs dataset, with a few exceptions. In the laquinimod dataset, a significantly greater FSS-Bowel and Bladder and smaller normalized brain volume (NBV) are observed (whereas these did not reach the same level of significance in the anti-CD20-Abs test set), and the difference in T25FW is not statistically significant (*p <* 0.05). A smaller NBV was found in the responder group, but this only reached significance at the 50th percentile threshold. Nonetheless, the direction of the effect for these differences is concordant between the two test sets.

## 3 Comparison to baseline models

The performance of the non-linear model described in this paper is compared to numerous other baseline models in Table 3, as measured by the *AD*_*wabc*_ on the anti-CD20-Abs test set and on the laquinimod dataset. The MLP outperforms all other baselines on this metric, but some models (such as a linear regression model with L2 regularization (ridge regression) and a CPH model) compare favorably on one of the two datasets. Without pre-training on the RRMS dataset, the performance of the MLP is still strong but inferior to the fine-tuned model.We also tested a prognostic MLP which is only trained to predict progression on placebo, and which uses this prediction in place of the CATE estimate (assumes that more rapid progression leads to greater potential for treatment effect). This model performs well, achieving the second best score on the anti-CD20-Abs dataset. All single feature models are inferior to the MLP and CPH models except for the T2 lesion volume / disease duration model which falls between the these two models in terms of performance on the anti-CD20-Abs test set.

**Table 3:** Comparison of model performance (measured by *AD*_*wabc*_) on the mixed dataset composed of ORATORIO and OLYMPUS (anti-CD20-Abs) and on the dataset composed of ARPEGGIO (laquinimod).

|  | Anti-CD20-Abs | Laquinimod |
| --- | --- | --- |
| **Single feature\*:** |  |  |
| Negative disease duration | 0.0225 | 0.0114 |
| Negative age | 0.0067 | -0.0287 |
| Negative EDSS | 0.0264 | 0.0074 |
| Negative 9HPT dominant hand | -0.0109 | 0.0023 |
| Negative 9HPT non-dominant hand | -0.0012 | -0.0006 |
| Negative T25FW | 0.0033 | 0.0020 |
| T2 lesion volume | 0.0167 | -0.0051 |
| Gad count | 0.0021 | NaN<sup>‡</sup> |
| **Feature / disease duration ratio<sup>†</sup>:** |  |  |
| Age / disease duration | 0.0268 | 0.0138 |
| EDSS / disease duration | 0.0021 | 0.0020 |
| 9HPT dominant hand / disease duration | 0.0238 | 0.0146 |
| 9HPT non-dominant hand / disease duration | 0.0179 | 0.0098 |
| T25FW / disease duration | 0.0257 | 0.0049 |
| T2 lesion volume / disease duration | 0.0432 | 0.0164 |
| Gad count / disease duration | 0.0030 | NaN<sup>‡</sup> |
| **Regression model using all features:** |  |  |
| MLP (our model) | 0.0565 | 0.0211 |
| MLP (no pre-training<sup>§</sup>) | 0.0486 | 0.019 |
| MLP (prognostic model<sup>¶</sup>) | 0.0408 | 0.0170 |
| Ridge Regression | 0.0227 | 0.0194 |
| **Survival model using all features:** |  |  |
| CPH | 0.0305 | 0.0031 |

\*The value of the feature is taken to be the CATE estimate for an individual. For example, the “T2 lesion volume” model uses the value of an individual’s T2 lesion volume as the CATE estimate for that individual, such that a larger baseline volume predicts a larger treatment effect. A “negative” feature implies that the CATE estimate is the negative of the value of the feature. For example, the “negative disease duration” model predicts a larger treatment effect with shorter disease duration.
<sup>†</sup>The value of the feature divided by the disease duration is taken to be the CATE estimate for an individual. For example, the “EDSS / disease duration” model predicts a larger treatment effect with a more rapid historical rate of change in the EDSS over time.
<sup>‡</sup>Value for $AD_{wabc}$ could not be computed due to low variance in values for Gad lesions in the laquinimod dataset.
<sup>§</sup>This MLP was trained without pre-training on the RRMS dataset.
<sup>¶</sup>The value of the predicted slope of disability progression on the placebo arm is used as the CATE estimate. In other words, a patient predicted to progress more rapidly on placebo (worse prognosis) predicts a larger treatment effect.
EDSS = Expanded Disability Status Scale; <sup>1</sup>FSS = Functional Systems Score; T25FW = timed 25-foot walk; 9HPT = 9-hole peg test; Gad = Gadolinium-enhancing lesion; MLP = Multi-layer perceptron.

In OLYMPUS, Hawker *et al*. [10] identify a cutoff of age < 51 years and gadolinium-enhancing (Gad) lesion count > 0 at baseline as predictive of treatment effect. Using their definition, 21.9% and 11.3% of the patients in the the anti-CD20-Abs and laquinimod datasets, respectively, would be classified as responders. This is more restrictive than our most restrictive threshold which selects the 30% predicted to be most responsive. The HR for these predicted responders is 0.91 (95% CI, 0.392-2.11; *p* = 0.831), and 0.305 (95% CI, 0.0558-1.67; *p* = 0.147) for the anti-CD20-Abs and the ARPEGGIO patients, respectively, indicating no improvement in treatment effect compared to the whole group.

Finally, we compared our approach to the traditional phase 2 approach which typically uses an MRI-based surrogate outcome (brain atrophy being the most common) which is thought to be correlated with the clinical outcome of interest but that is more sensitive to the underlying biological processes or that has a lower variance, in order to increase a study’s statistical power. For example, suppose our anti-CD20-Abs test set (*n* = 297) was a small phase 2 trial testing anti-CD20-Abs with brain atrophy as the primary outcome. Measuring brain atrophy at the 48 week MRI for the anti-CD20-Abs, the mean difference between the treatment arms is 0.066 (95% CI, -0.397 to 0.529; *p* = 0.7786). Looking at ORATORIO patents separately, since ORATORIO was the only positive trial in the anti-CD20-Abs dataset, the mean difference is 0.110 (95% CI, -0.352 to 0.572; *p* = 0.6379). Brain atrophy would therefore not have been able to detect a significant effect for ocrelizumab or for anti-CD20-Abs.

### 3.1 Simulating a phase 2 clinical trial enriched with predicted responders

To understand the effect of enriching a future clinical trial studying novel B-cell depleting agents, we simulated both a one and a two-year randomized clinical trial using populations enriched with predicted responders. Using our model to predict sub-groups of responders to anti-CD20-Abs across a variety of thresholds, we can calculate the 1-year CDP24 event rate and 1-year HR for these sub-groups, which can then be used for sample size estimation. Table 4 shows the sample size needed detect a significant difference across various degrees of enrichment.

**Table 4:**
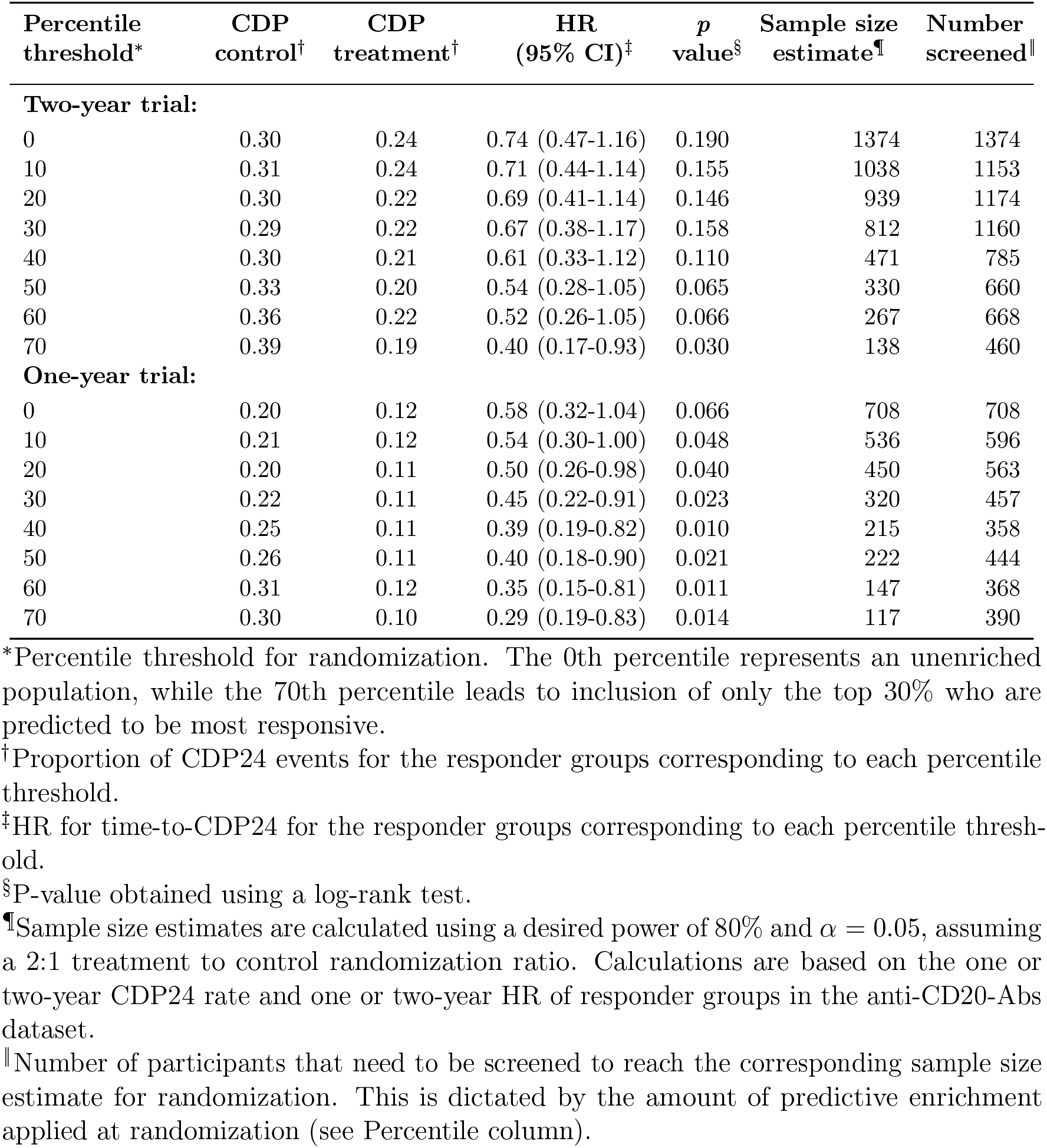
Estimated sample size for a one or two-year placebo-controlled randomized clinical trial of anti-CD20-Abs, using different degrees of predictive enrichment.

Using the 50th percentile as a threshold for randomization in a 1-year long trial as an example, a total of 444 individuals would be screened and the top 50% who are predicted to be most responsive would be randomized (*n* = 222). This leads to a 3-fold reduction in the number of patients that need to be randomized while screening 1.6 times less patients compared to the scenario where all participants are randomized into a one-year study (*n* = 708).

## 4 Discussion

This work addresses the lack of a sufficiently predictive biomarker of treatment response, which has hampered progress in the field of PPMS by preventing efficient phase 2 clinical trials. We describe a deep learning solution to increasing the efficiency of early proof-of-concept clinical trials based on a multi-headed MLP architecture designed for CATE estimation. This approach can consistently identify and rank treatment effect among patients exposed to anti-CD20-Abs, and could reduce by several fold the sample size required to detect an effect in a short one or two-year long trial. We validate our model using a dataset composed of patients exposed to anti-CD20-Abs, and a second dataset of patients exposed to laquinimod. We demonstrate that a model trained to predict response to anti-CD20-Abs can also generalize to laquinimod, a medication with a very different mechanism of action, suggesting that there exists disease-agnostic predictors of response.

The model’s predicted responders were enriched in numerous baseline features, including a younger age, shorter time from symptom onset, higher disability scores, and more lesion activity. Similarly, in subgroup analyses from OLYMPUS, an age less than 51 years and presence of Gad lesions at baseline was also found to be associated with increased response [10]. Signori *et al*. [14] also found that younger age and the presence of Gad were associated with greater treatment effect in RRMS. In a study by Bovis *et al*. [2], a response scoring function obtained via CPH models in RRMS also identified Gad lesions and a higher normalized brain volume as predictive of treatment effect, although older age was found to be more predictive in the combination they studied.

In our experiments, a non-linear model (MLP) outperformed other linear (and log-linear) baselines, suggesting that complex relationships exist between the baseline features and treatment effect. Nonetheless, a prognostic model (that predicts response to a medication solely based on the prediction of progression on placebo) also performed well, suggesting that poor prognosis is also predictive of treatment effect. A prognostic model could therefore be helpful in cases where drugs with very different mechanisms of action (e.g. targetting remyelination, or neurodegeneration) are being tested, in which case a model trained to predict treatment effect on an anti-inflammatory drug might perform less well than a prognostic model.

Interestingly, despite a balanced dataset with respect to gender, our model was better at identifying responders in women compared to men. We also noted that the model performed better in individuals >= 51, disease duration < 5 years, and/or an EDSS < 4.5. These findings suggest further studies are needed to determine whether and why predictors of response might differ depending on the stage of disease and sex.

Predictive enrichment is not the only approach to increase the efficiency of clinical trials in PPMS. However, the traditional approach of using a potential surrogate marker (such as brain atrophy) as part of a phase 2 study did not succeed in identifying a significant effect in our experiments, and may therefore limit early identification of effective therapies. While frequently used in phase 2 trials as a primary outcome, several studies on PPMS [15], RRMS [16], and secondary progressive multiple sclerosis (SPMS) [17] suggest no to modest correlation with clinical disability progression based on EDSS even after four to eight years of follow-up.

Another strategy has been to infer from a positive RRMS trial that a drug might be effective on disability progression. Two notable examples are ocrelizumab and siponimod, which were first found to be efficacious in the RRMS population in OPERA I/II [7] and BOLD [18], respectively, before being tested in the PPMS trial ORATORIO [9] and the SPMS trial EXPAND [19], respectively. From a predictive enrichment standpoint, baseline T2 lesion burden has been found to correlate with future disability and disability progression, at least modestly [20–23]. Evidence is less robust for Gad lesions, since some authors [24] have demonstrated modest correlations with future disability at least 2 years from baseline, while others [22] have not. In our experiments, a treatment effect estimation model based on either Gad count or T2 lesion volume alone performed poorly. Only the rate of accumulation of T2 lesions over time (measured from the time of symptom onset) was predictive. Even if the inflammatory hypothesis was correct, a predictive enrichment strategy is likely to be more efficient than awaiting the results of a RRMS study testing the same drug, particularly given that the power of a follow-up PPMS study is likely to be insufficient, as shown by the small proportion of responders to anti-CD20-Abs in our experiments, the dramatic difference in effect size between the inflammatory and progression-related outcomes, and the numerous examples of effective drugs for RRMS that had no identifiable effect on slowing disability progression in progressive multiple sclerosis (PMS) [10, 11, 25–29].

Finally, the Food and Drug Administration has published a guidance document with suggestions regarding the design of predictively enriched studies [30]. One approach might be to first conduct a small trial of a short duration as a proof of concept in patients predicted to be highly responsive. If a significant effect is detected, a larger/longer followup study with a more inclusive (less enriched) population can be attempted with more confidence. It is also possible that, on the basis of a strong effect in the enriched responder group, the proof of concept would be sufficient for drug approval to be granted for the un-enriched population, given the significant unmet need and irreversible consequences of disability progression. To limit the risk that the predictive model is found to be inaccurate on the study population, stratified randomization can be used by having two parallel groups: the primary group (which would be adequately powered to detect an effect) would be an enriched responder group, while the secondary group would randomize predicted non-responders. Although the non-responder group would not be powered to detect an effect, it would provide a rough estimate of the effectiveness of the drug in this group and help guide design decisions for follow-up trials. The two groups could also be merged in a pre-planned analysis, to provide an estimate of the effect in the combined population.

Limitations of this work include the choice of model. Interpretability of black-box algorithms such as neural networks (reviewed elsewhere [31]) remains an area of active research. Although our MLP outperformed linear baselines, MLPs are more difficult to train and at higher risk of overfitting. Moreover, we made heavy use of several regularization schemes to prevent this. Our hyperparameter tuning procedure is also one of many that can be designed. We used MRI-derived lesion and volumetric measures computed during the individual clinical trials, which could potentially ignore more subtle predictive features found within the MRI voxel-level data. Learning these features in a data-driven fashion through convolutional neural networks is the subject of ongoing work, but this can easily be appended to our MLP architecture. More data is needed from drugs with diverse mechanisms of action to fully grasp the extent to which predictors of anti-inflammatory drugs are applicable to other drug classes, including neurodegenerative targets. Finally, it remains unknown if patients for whom our model predicted minimal effect over two to four years could benefit after longer periods of administration. Answering this question would require longer-term observational data.

## 5 Online Methods

### 5.1 Data

Data is taken from six different randomized clinical trials (*n* = 3, 830): OPERA I [7], OPERA II [7], BRAVO [8], ORATORIO (NCT01194570) [9], OLYMPUS (NCT00087529) [10], and ARPEGGIO [11] (ClinicalTrials.gov numbers, NCT01247324, NCT01412333, NCT00605215, NCT01194570, NCT00087529, NCT02284568, respectively). All three trials enrolled adults with PPMS and had similar inclusion criteria (see the original publications for details). We excluded participants who spent less than 24 weeks in the trial, who had less than two clinical visits, or who were missing one or more input features at the baseline visit. Therefore, it is important to appreciate that the data included in our work are not an exact reproduction of those used in the clinical trials.

All clinical/demographic and MRI features that were consistently recorded as part of all 6 clinical trials (total of 19 features) were used to train our model. Values were recorded at the baseline visit (immediately before randomized treatment allocation), and are a combination of binary (sex), ordinal (EDSS, FSS), discrete (Gad count), and continuous variables (age, height, weight, disease duration, T25FW, 9-hole peg test (9HPT), T2 lesion volume, Gad count, and NBV). Disease duration was estimated from the time of symptom onset.

Lesion segmentation and volumetric measurements were done according to the individual study’s methodology. To account for different segmentation algorithms with different sensitivities, the subset of the samples that fulfilled the intersection of the inclusion criteria for all trials were used to compute a scaling factor for all MRI metrics such that their scaled range from -3S to +3SD matches that of a reference trial in the training set (ORATORIO for the PPMS trials, and OPERA I/II for the RRMS trials). Once computed, the scaling factors were applied to all samples.

The following right-skewed distributions were log-transformed: NBV, T2 lesion volume, T25FW, and 9HPT. Gad counts were binned into bins of 0, 1, 2, 3, 4, 5-6, 7-9, 10-14, 15-19 and 20+ lesions. Finally, to improve convergence during gradient descent, all non-binary features were standardized by subtracting the mean and dividing by the standard deviation, both calculated from the training dataset [32].

### 5.2 Outcome definition

The primary outcome used in clinical trials assessing the efficacy of therapeutic agents on disease progression is the time to confirmed disability progression (CDP) at 12, or 24 weeks. We use CDP24 because it is a more robust indication that disability accrual will be maintained after 5 years [33]. CDP24 is most commonly based on the EDSS, a scale going from 0 (no disability) to 10 (death), in discrete 0.5 increments (except for a 1.0 increment between 0.0 and 1.0). A CDP24 event is defined as a 24-week sustained increase in the EDSS of 0.5 for baseline EDSS values *>* 5.5, of 1.5 for a baseline EDSS of 0, and of 1.0 for EDSS values in between. This difference in the increment required to confirm disability progression is commonly adopted in clinical trials, and partially accounts for the finding that patients transition through the EDSS scores at different rates [34].

While it is possible to predict time-to-event using traditional machine learning methods if workarounds are used to address right-censored data or using machine learning frameworks specifically developed to model survival data (reviewed elsewhere [35]), we chose not to model time-to-CDP24 because of limitations inherent in this metric. As outlined by Healy *et al*. [36], CDP reflects not only the rate of progression but also the baseline stage of the disease, which is problematic because the stage is represented by a discretized EDSS at a single baseline visit. This results in a noisy outcome label which could make it harder for a model to learn a representation that relates to the progressive biology which we are trying to model.

We therefore model the rate of progression directly by fitting a linear regression model onto the EDSS values of each individual participant over multiple visits (see Supplementary Methods 2 for details) and take its slope to be the outcome label that our MLP uses for training. One advantage of the slope outcome over time-to-CDP24 is that it can be modeled using any type of regression model. We revert to using time-to-CDP24 for model evaluation to facilitate comparison with treatment effect survival metrics reported in the original clinical trial publications.

### 5.3 Treatment effect modeling

To enrich clinical trials with individuals predicted to have an increased response to treatment, it is helpful to begin with the definition of individual treatment effect (ITE) according to the Neyman/Rubin Potential Outcome Framework [37]. Let the ITE for individual *i* be *τ*_*i*_, then

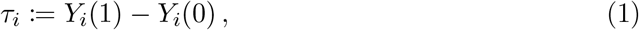

where *Y*_*i*_(1) and *Y*_*i*_(0) represent the outcome of individual *i* when given treatment and control medications, respectively. The *Fundamental Problem of Causal Inference* [38] states that the ITE is unobservable because only one of the two outcomes is realized in any given patient, dictated by their treatment allocation. *Y*_*i*_(1) and *Y*_*i*_(0) are therefore termed *potential* outcomes or, alternatively, factual (observed) and counterfactual (not observed) outcomes.

Ground-truth can nevertheless be observed at the group level in specific situations, such as randomized control trials, because treatment allocation is independent of the outcome. We provide a detailed discussion of two important estimands, the average treatment effect (ATE) and the CATE in Supplementary Methods 1. Briefly, ATE represents the average effect when considering the entire population, while CATE considers a sub-population characterized by certain characteristics (e.g. 40 year-old women with 2 Gad lesions at baseline). We use CATE estimation to frame the problem of predicting treatment response for individuals.

The best estimator for CATE is conveniently also the best estimator for the ITE in terms of mean squared error (MSE) [5]. Several frameworks have been developed to model CATE, but a simple metalearning approach which decomposes the estimation into sub-tasks that can be solved using any supervised machine learning model provides a flexible starting point [5]. For a broader survey of methods, see the survey on uplift modeling by Gutierrez & Gérardy [3] (the uplift literature has contributed extensively to the field of causal inference, particularly when dealing with randomized experiments from an econometrics perspective).

In this work, an MLP was selected as the base model due to its high expressive power and flexibility to be integrated into larger end-to-end-trainable neural networks consisting of different modules (such as convolutional neural networks). We used a multi-headed architecture, with a common trunk and two output heads: one for modeling the potential outcome on treatment, 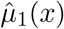, and the other to model the potential outcome on placebo, 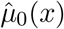. For inference, the CATE estimate 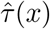 given a feature vector *x* can be computed as:

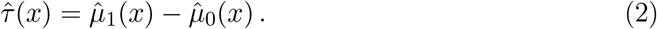

We use 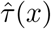 as the predicted treatment effect for an individual with characteristics *x*. Note that we multiplied all 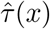 values by -1 in this paper to simplify interpretation in Section 2 (Results), such that a positive effect indicates improvement, while a negative effect indicates worsening on treatment.

This multi-headed approach can be seen as a variant of the T-Learner described by Künzel *et al*. [5], except that the two base models in our case share weights in the common trunk. Our network is similar to that conceptualized by Alaa *et al*. [39], but without the propensity network used to correct for any conditional dependence between the treatment allocation and the outcome given the input features, since our dataset comes from randomized data.

To decrease the size of the hyperparameter search space, we fixed the number of layers and only tuned the layer width. We used one common hidden layer and one treatment-specific hidden layer. Additional common or treatment-specific layers could be used if necessary, but given the low dimensionality of our feature-space and the relatively small sample size, the network’s depth was kept small to avoid over-fitting. The inductive bias behind our choice of using a multi-headed architecture is that disability progression can have both disease-specific and treatment-specific predictors of disability progression, which can be encoded into the common and treatment-specific hidden layer representations, respectively. Consequently, the common hidden layers can learn from all the available data, irrespective of treatment allocation. Rectified linear unit (ReLU) activation functions were used at hidden layers for non-linearity.

### 5.4 Training

The model was trained in two phases, depicted in Fig. 3. In the first phase, a 5-headed MLP was pre-trained on an RRMS dataset to predict the slope outcome on each treatment arm. In the second phase, the parameters of the common layers were frozen, and the output heads were replaced with two new randomly initialized output heads for fine-tuning on the PPMS dataset to predict the same outcome.

**Figure 3:**
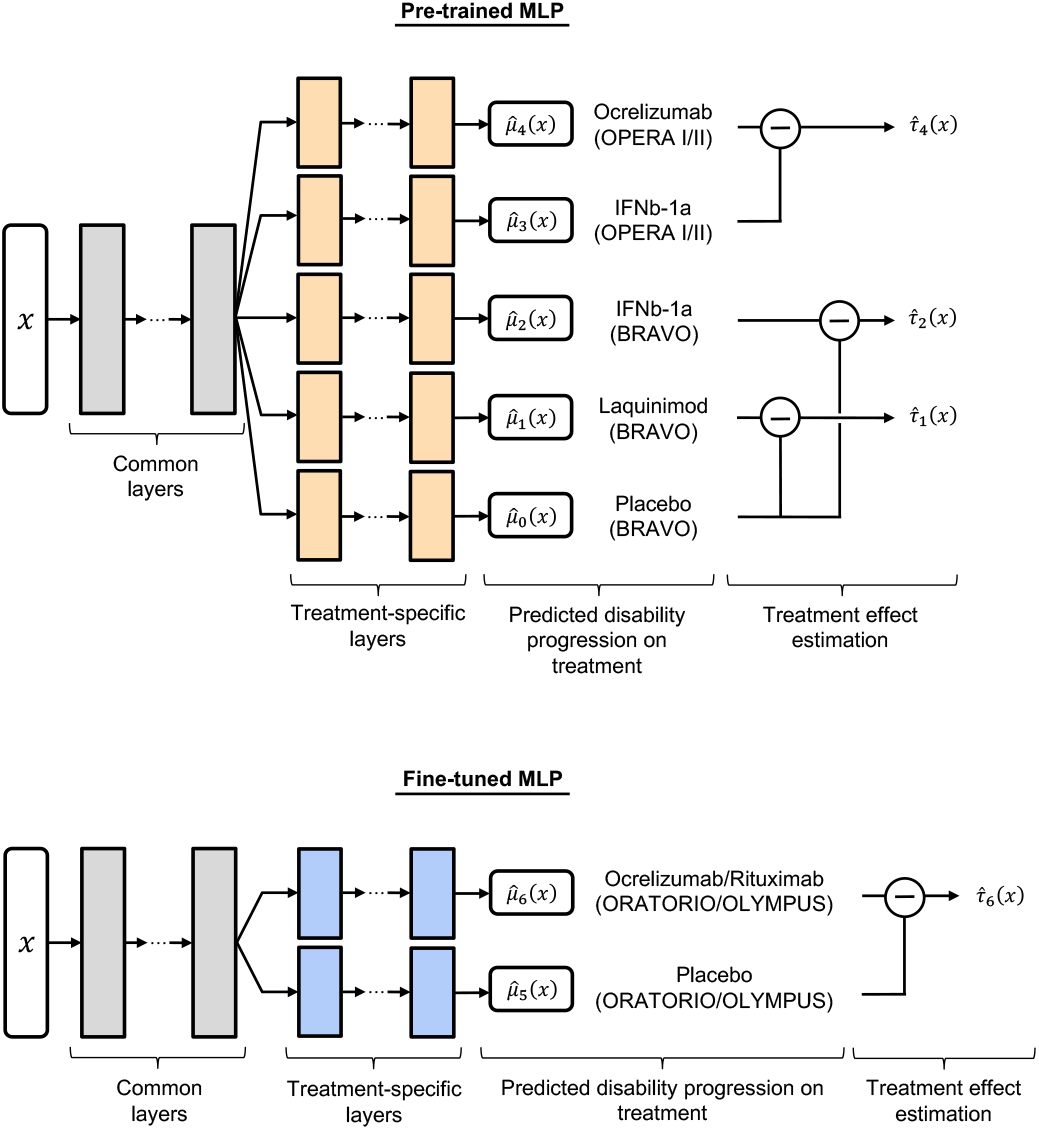
Multi-headed multilayer perceptron (MLP) architecture for CATE estimation. The MLP was first pre-trained on a relapsing-remitting multiple sclerosis dataset (top), followed by fine tuning on a primary progressive multiple sclerosis dataset (bottom). Subtraction symbols indicate which treatment and control are being subtracted for the CATE estimate. Grey-colored layers indicate the common layers that are transferred from the pre-trained MLP to the fine-tuning MLP, at which point their parameters are frozen and only the parameters of the blue-colored layers are updated. The orange-colored layers are discarded after the pre-training step. 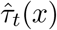: CATE estimate for treatment *t* given feature vector *x*. 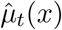: predicted potential outcome on treatment *t*. IFNb-1a = Interferon beta-1a.

Optimization was done using mini-batch gradient descent with momentum. To prevent overfitting, the validation loss was monitored during 4-fold cross-validation (CV) to early-stop model training at the epoch with the lowest MSE, up to a maximum of 100 epochs. Dropout and L2 regularization were used, along with a max-norm constraint on the weights [40], to further prevent overfitting.

Mini-batches were sampled in a stratified fashion to preserve the proportions of participants receiving active treatment and placebo. Backpropagation was done using the MSE calculated at the output head that corresponds to the treatment that the patient was allocated to, *t*_*i*_ (the output head with available ground-truth). The squared errors from each output head were then weighted by *n*_*s*_*/*(*m * n*_*t*_), where *n*_*s*_ represents the total number of participants in the training split, *n*_*t*_ represents the number of participants in the treatment arm corresponding to the output head of interest, and *m* represents the total number of treatment arms. This compensates for treatment allocation imbalance in the dataset.

We aimed to reduce variance by using the early-stopped models obtained from each CV fold as members of an ensemble. This ensemble’s prediction is the mean of its members’ predictions, and is used for inference on the unseen test set.

A random search was used to identify the hyperparameters with the best validation performance (learning rate, momentum, L2 regularization coefficient, hidden layer width, max norm, dropout probability). We used CV aggregation, or cross-validation aggregation (crogging) [41], to improve the generalization error estimate using our validation metrics. Crogging involves aggregating all validation set predictions (rather than the validation metrics) and computing one validation metric for the entire CV procedure. The best model during hyperparameter tuning was selected during CV on the basis of two validation metrics: the MSE of the factual predictions, and the *AD*_*wabc*_ (described in detail in Supplementary Methods 3). We combine both validation metrics during hyperparameter tuning by choosing the model with the highest *AD*_*wabc*_among all models that fall within 1 SD of the best performing model based on the MSE loss. The SD of the best performing model’s MSE is calculated from the loss values obtained in the individual CV folds.

### 5.5 Baseline models

The performance of the multi-headed MLP was compared to ridge regression and CPH models. Both models were used as part of a T-learner configuration (as defined by Künzel *et al*. [5]). Hyperparameter tuning was done on the same folds and with the same metrics as for the MLP.

### 5.6 Statistical Analysis

Hazard ratios were calculated using CPH models and associated p-values from log-rank tests.

### 5.7 Software

All experiments were implemented in Python 3.8 [42]. MLPs were implemented using the Pytorch library [43]. Scikit-Learn [44] was used for the implementation of ridge regression, while Lifelines [45] was used for CPH. For reproducibility, the same random seed was used for data splitting and model initialization across all experiments.

### 5.8 Data Availability

Data used in this work are controlled by pharmaceutical companies and therefore are not publicly available. Access requests should be forwarded to data controllers via the corresponding author.

## 6 Acknowledgments

The authors are grateful to the International Progressive MS Alliance for supporting this work and to the companies who generously provided the clinical trial data that made it possible: Biogen, BioMS, MedDay, Novartis, Roche / Genentech, and Teva.

## 7 Funding

This investigation was supported (in part) by an award from the International Progressive Multiple Sclerosis Alliance (award reference number PA-1412-02420), by an endMS Personnel Award from the Multiple Sclerosis Society of Canada (Falet, JR), and by a Canada Graduate Scholarship-Masters Award from the Canadian Institutes of Health Research (Falet, JR). Falet, JR is also being supported through the Fonds de recherche du Québec - Santé / Ministère de la Santé et des Services sociaux training program for specialty medicine residents with an interest in pursuing a research career, Phase 1.

## 8 Competing Interests

Arnold, DL, reports consulting fees from Albert Charitable Trust, Alexion Pharma, Biogen, Celgene, Frequency Therapeutics, Genentech, Med-Ex Learning, Merck, Novartis, Population Council, Receptos, Roche, and Sanofi-Aventis, grants from Biogen, Immunotec and Novartis, and an equity interest in NeuroRx. Sormani, MP, has received personal compensation for consulting services and for speaking activities from Merck, Teva, Novartis, Roche, Sanofi Genzyme, Medday, GeNeuro, and Biogen. Precup, D, works part-time for DeepMind. The remaining authors report no competing interests.

## Notes

### Author Declarations

Ethics committee of the Montreal Neurological Institute gave ethical approval for this work.

