## Supplementary for "Estimating treatment effect for individuals with progressive multiple sclerosis using deep learning"

### Supplementary Information

### Supplementary Methods

#### 1 Treatment Effect Estimation

To enrich clinical trials with individuals predicted to have an increased response to treatment, it is helpful to begin with the definition of individual treatment effect (ITE) according to the Neyman/Rubin Potential Outcome Framework [1]. Let the ITE for individual $i$ be $\tau_i$, then

$$\tau_i \coloneqq Y_i(1) - Y_i(0)\,, \tag{1}$$

where $Y_i(1)$ and $Y_i(0)$ represent the outcome of individual $i$ when given treatment and control medications, respectively. The *Fundamental Problem of Causal Inference* [2] states that the ITE is unobservable because only one of the two outcomes is realized in any given patient, dictated by their treatment allocation. $Y_i(1)$ and $Y_i(0)$ are therefore termed *potential* outcomes or, alternatively, factual (observed) and counterfactual (not observed) outcomes.

Ground-truth can nonetheless be observed at the group level. The average treatment effect (ATE) is defined as the expected difference between both potential outcomes:

$$ATE \coloneqq \mathbb{E}[Y(1) - Y(0)] = \mathbb{E}[Y(1)] - \mathbb{E}[Y(0)]\,. \tag{2}$$

Equation 2 is still in terms of unobservable causal quantities, so additional assumptions are needed. While a detailed discussion of the underlying assumptions is beyond the scope of this paper, in specific situations, such as randomized control trials, where the outcome is independent of treatment allocation, the ATE can identified from the observed outcome $Y$ as follows

$$\mathbb{E}[Y|T = 1] - \mathbb{E}[Y|T = 0]\,, \tag{3}$$

where $T \in \{0, 1\}$ is the treatment allocation. Broadly speaking, the ATE (sometimes formulated as a ratio instead of a difference) is what is estimated in clinical trials, but here we seek to estimate the ATE of a sub-group of patients conditioned on their baseline characteristics, a d-dimensional feature vector $x \in \mathcal{X} \subseteq \mathbb{R}^d$. The conditional average treatment effect (CATE), denoted $\tau(x)$, is defined as:

$$\tau(x) \coloneqq \mathbb{E}[Y(1)|X = x] - \mathbb{E}[Y(0)|X = x]\,, \tag{4}$$

which can similarly be rewritten in terms of the observed outcome $Y$ in the context of randomized controlled trials, where $\{(Y(0), Y(1)) \perp\!\!\!\perp T\}|X$:

$$\tau(x) = \mathbb{E}[Y|X = x, T = 1] - \mathbb{E}[Y|X = x, T = 0] = \mu_1(x) - \mu_0(x)\,. \tag{5}$$

A CATE estimator, $\hat{\tau}(x) = \hat{\mu}_1(x) - \hat{\mu}_0(x)$, can be parametrized by a neural network trained on an observational dataset $\mathcal{D} = \{(x_i, y_i, t_i)\}_{i=1}^n$. In this paper, we learn a multi-headed multilayer perceptron (MLP) in which $\hat{\mu}_1(x)$ and $\hat{\mu}_0(x)$ share parameters in the earlier layers but have distinct parameters in the output heads. We use $\hat{\tau}(x_i)$ as the estimate for the treatment effect of an individual, $\hat{\tau}_i$.

#### 2 Slope Outcome

We assume that progression is slow over the course of the one to two year duration of a phase 2 or 3 clinical trial such that the Expanded Disability Status Scale (EDSS) value at time $t$ following treatment initiation can be modeled as the linear relationship

$$EDSS = \beta_0 + \beta_1 t\,, \tag{6}$$

where $\beta_0$ and $\beta_1$ are the regression coefficients. Using the method of ordinary least squares for linear regression, estimates $\hat{\beta}_0$ and $\hat{\beta}_1$ are found using all available timepoints $t$. Each patient $i$ has a separate slope of disability progression, $\hat{\beta}_{1,i}$, found by fitting a linear regression model to their own EDSS values. This slope is then used as the ground-truth outcome $y_i$ that we train a neural network to predict:

$$y_i = \hat{\beta}_{1,i}\,. \tag{7}$$

To compute the slope, a minimum of two timepoints $t$ must be available for each patient. We also require that the duration between the first and last timepoints be greater than 24 weeks, given that we are evaluating our model's performance using 24-week confirmed disability progression (CDP24). Participants who do not fulfill these two requirements are excluded from the dataset. The average number of visits used to compute the slopes was 12.23 (SD 2.86; range 3-24).

Note that the definition of confirmed disability progression (CDP) used in clinical trials depends on the baseline EDSS of the individual. For a CDP event to occur, a participant who has a baseline EDSS of 0 requires an increase in EDSS of 1.5, while a baseline of $> 5.5$ requires an increase of 0.5. Baseline values in between require an increase of 1.0. Therefore, in order for our slope outcome to closely resemble the changes in EDSS that are required to reach CDP, we scaled the EDSS values prior to fitting the linear regression models, such that the increase necessary for a CDP event to occur approximately maps to an increase of 1.0 after the scaled transform:

$$f(EDSS) = \begin{cases} \frac{EDSS}{1.5}, & \text{if } EDSS \leq 1.5 \\ EDSS - 0.5, & \text{if } 1.5 < EDSS \leq 6.0 \\ \frac{EDSS-6.0}{0.5} + 5.5, & \text{if } EDSS > 6.0 \end{cases} \tag{8}$$

We use the scaled values, $f(EDSS)$ in place of the EDSS when fitting the linear regression model. $f(EDSS)$ is plotted in Supplementary Fig. 3.

#### 3 Weighted Average Treatment Difference Curve

Following Zhao *et al.* [3], we define a conditional expectation, $AD(c)$, which reflects the ATE of a sub-group of patients who are predicted by our model to have a treatment effect greater than a threshold value $c$:

$$AD(c) = \mathbb{E}[\,Y(1) - Y(0)\,|\,\hat{\tau}_i \geq c\,]\,. \tag{9}$$

The conditional expectation for $Y(1) - Y(0)$ is estimated using the restricted mean survival time (RMST) for the time-to-CDP24, truncated at 2 years [4]. By defining the conditional expectation in terms of the RMST instead of the slope outcome used as the target for training the neural network, the $AD(c)$ better reflects how well our model can identify responders using a survival-based metric, which is ultimately what clinical trials will use.

The $AD(c)$ behaves as a population selector for predictive enrichment, whereby patients expected to respond with effect size greater than a desirable threshold $c$ can be enrolled in a clinical trial or recommended the medication in a clinical setting.

If patients are ranked accurately according to their predicted responsiveness to the active medication, then the resultant $AD(c)$ curve should have a large area under the curve, $AD_{auc}$. The $AD_{auc}$ is therefore a useful evaluation metric. We compute the $AD_{auc}$ using polygon approximation with operating points every 10 percentiles from 0 until the 70th percentile for better computational efficiency, while we use 1 percentile increments for reporting test metrics and for visualization purposes in this paper. Following Zhao *et al.* [3], we then subtract the effect size of the entire (unenriched) population from the $AD_{auc}$ to facilitate the comparison of different models. This metric is called the area between curves, or $AD_{abc}$, and can be written as

$$AD_{abc} = AD_{auc} - AD(\hat{\tau}_{(0)})\,, \tag{10}$$

where $\hat{\tau}_{(0)}$ represents the minimum predicted treatment effect in the evaluation set. We further weigh the $AD_{abc}$ by multiplying it to a measure of monotonicity to promote a monotonically increasing $AD(c)$, since monotonicity indicates that the model can rank response accurately throughout the range of possible responsiveness. To do so, we use the Spearman's rank correlation coefficient, $\rho$, calculated between the $AD_{abc}$ values and the thresholds $c$, as the scaling factor for the $AD_{abc}$:

$$AD_{wabc} = \rho AD_{abc}\,. \tag{11}$$

#### Supplementary Figures

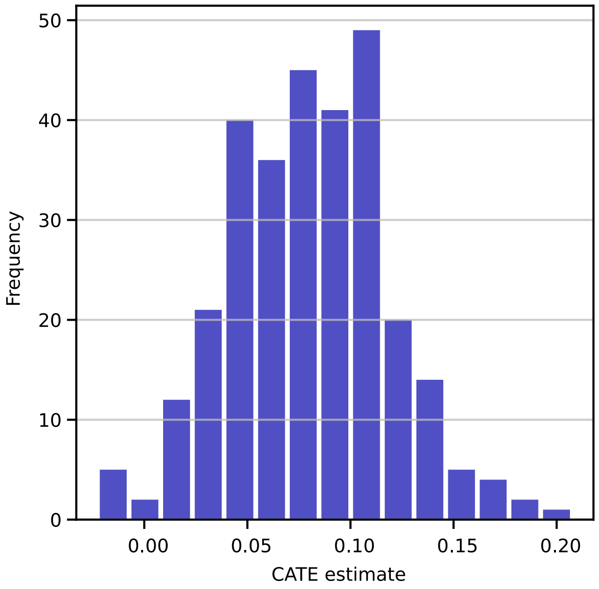


Supplementary Figure 1: Histogram of CATE estimates for the anti-CD20-Ab test set. Positive numbers indicate a predicted benefit from anti-CD20-Abs over placebo, 0 indicates no predicted benefit, and negative numbers indicate predicted harm.

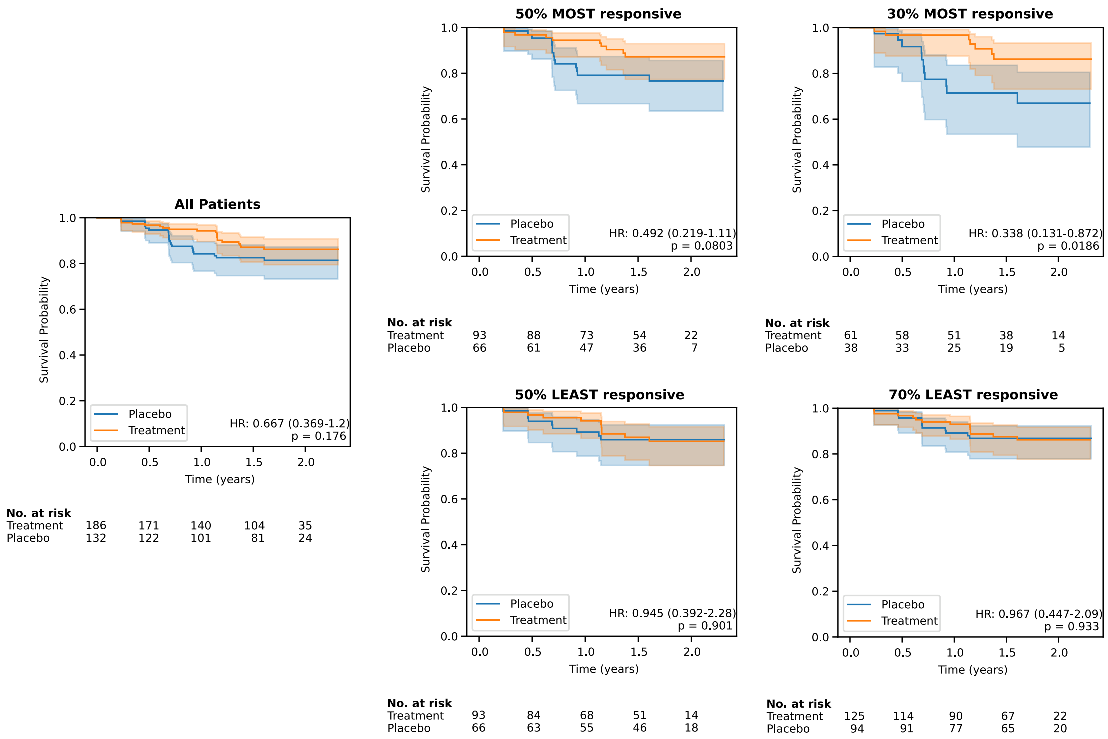


Supplementary Figure 2: Kaplan-Meyer curves for predicted responders and non-responders to laquinimod, defined at two thresholds of predicted effect size. These are compared to the whole group (left). Survival probability is measured in terms of time-to-CDP24 using the EDSS. $p$ values are calculated using log-rank tests. Kaplan-Meyer curve 95% confidence intervals are estimated using Greenwood's Exponential formula.

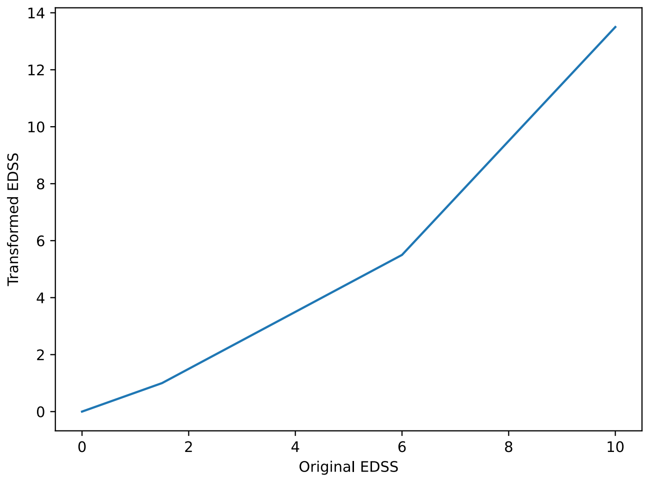


Supplementary Figure 3: Expanded Disability Status Scale transformation to account for the baseline-dependent definition of confirmed disability progression.

### Supplementary Tables

Supplementary Table 1: Feature features and outcomes per treatment arm for the relapsing-remitting pre-training dataset.

| | **Ocrelizumab** | | **IFNb-1a SC** | | **IFNb-1a IM** | **Laquinimod** | **Placebo** |
|---|---|---|---|---|---|---|---|
| | OPERA I | OPERA II | OPERA I | OPERA II | BRAVO | BRAVO | BRAVO |
| | n=320 | n=335 | n=295 | n=329 | n=412 | n=407 | n=422 |
| **Demographics:** | | | | | | | |
| Age (years) | 37.35 (9.36) | 37.44 (8.93) | 37.25 (9.54) | 37.39 (8.82) | 38.02 (9.41) | 37.02 (9.19) | 37.50 (9.59) |
| Sex (% male) | 35.00 | 37.01 | 32.20 | 31.61 | 32.28 | 35.63 | 28.67 |
| Height (cm) | 169.58 (8.91) | 169.59 (9.52) | 169.40 (9.18) | 168.66 (8.81) | 168.32 (8.57) | 169.12 (8.66) | 169.05 (8.64) |
| Weight (kg) | 74.31 (17.42) | 76.51 (16.97) | 75.25 (17.04) | 74.99 (19.00) | 69.63 (15.93) | 69.50 (15.04) | 69.52 (13.66) |
| Disease duration (years) | 6.71 (6.45) | 6.58 (5.95) | 6.08 (5.79) | 6.80 (6.28) | 6.93 (5.81) | 6.50 (5.80) | 6.90 (6.53) |
| **Disability Scores:** | | | | | | | |
| EDSS | 2.79 (1.22) | 2.68 (1.32) | 2.60 (1.26) | 2.74 (1.40) | 2.63 (1.15) | 2.65 (1.24) | 2.73 (1.18) |
| FSS-Bowel and Bladder | 0.56 (0.73) | 0.64 (0.79) | 0.60 (0.79) | 0.61 (0.81) | 0.52 (0.71) | 0.57 (0.76) | 0.54 (0.71) |
| FSS-Brainstem | 0.59 (0.81) | 0.48 (0.76) | 0.57 (0.77) | 0.50 (0.79) | 0.73 (0.78) | 0.78 (0.81) | 0.83 (0.82) |
| FSS-Cerebellar | 1.15 (1.02) | 1.03 (1.01) | 1.00 (0.96) | 1.04 (1.01) | 1.20 (0.96) | 1.21 (1.04) | 1.25 (0.99) |
| FSS-Cerebral | 0.50 (0.72) | 0.60 (0.81) | 0.55 (0.77) | 0.65 (0.83) | 0.64 (0.76) | 0.66 (0.74) | 0.70 (0.79) |
| FSS-Pyramidal | 1.71 (1.02) | 1.65 (1.05) | 1.54 (1.01) | 1.54 (1.05) | 1.79 (0.96) | 1.73 (1.00) | 1.75 (0.98) |
| FSS-Sensory | 1.17 (1.00) | 1.01 (1.00) | 1.04 (0.96) | 1.10 (1.01) | 0.94 (1.02) | 1.04 (1.04) | 1.02 (0.99) |
| FSS-Visual | 0.67 (0.84) | 0.68 (0.89) | 0.72 (0.88) | 0.69 (0.91) | 0.80 (1.09) | 0.79 (1.17) | 0.85 (1.25) |
| Mean T25FW (sec) | 7.80 (7.56) | 8.19 (11.83) | 7.04 (7.14) | 7.29 (7.64) | 6.31 (5.45) | 6.00 (2.89) | 6.04 (3.05) |
| Mean 9HPT dominant hand (sec) | 24.47 (17.66) | 23.80 (9.09) | 23.77 (17.37) | 24.52 (13.34) | 21.73 (5.87) | 21.98 (7.18) | 22.83 (17.16) |
| Mean 9HPT non-dominant hand (sec) | 26.85 (23.72) | 25.26 (13.02) | 24.51 (8.09) | 26.31 (19.01) | 23.13 (6.00) | 23.06 (6.86) | 23.87 (12.46) |
| **MRI metrics:** | | | | | | | |
| Gad count | 1.76 (4.49) | 1.81 (4.51) | 1.74 (4.93) | 1.96 (5.16) | 1.85 (6.86) | 1.84 (5.22) | 1.47 (5.88) |
| T2 Lesion Volume (mL) | 10.59 (14.25) | 11.28 (15.00) | 8.69 (10.13) | 10.19 (12.07) | 8.86 (10.55) | 9.69 (10.38) | 7.99 (8.95) |
| Normalized brain volume (L) | 1.50 (0.08) | 1.50 (0.09) | 1.50 (0.09) | 1.50 (0.09) | 1.59 (0.08) | 1.58 (0.10) | 1.59 (0.09) |
| **Outcome:** | | | | | | | |
| Slope (EDSS change / yr)* | -0.01 (0.39) | 0.00 (0.58) | 0.07 (0.47) | 0.09 (0.57) | 0.06 (0.72) | 0.04 (0.53) | 0.14 (0.83) |
| RMST (at 2 years)† | 1.97 | 1.95 | 1.93 | 1.92 | 1.93 | 1.93 | 1.90 |

Values in brackets are standard deviations, unless otherwise specified.

* Slope is based on the coefficient of regression from a linear regression model that is fit on an individual's EDSS values over time, as described in Section 5.2.

† RMST calculated at 2 years using time to 24-week confirmed disability progression on the EDSS.

RMST=Restricted mean survival time; IFNb-1a = Interferon beta-1a; IM = intramuscular; SC = subcutaneous; EDSS = Expanded Disability Status Scale; FSS = Functional Systems Score; T25FW = timed 25-foot walk; 9HPT = 9-hole peg test; Gad = Gadolinium-enhancing lesion.

Supplementary Table 2: Group statistics for predicted responders and non-responders to laquinimod at the 50th and 70th percentile thresholds.

| | **50th percentile threshold*** | | | | **70th percentile threshold*** | | | |
|---|---|---|---|---|---|---|---|---|
| | **Responders** | **Non-responders** | **Effect size (95% CI)†** | ***p* value‡** | **Responders** | **Non-responders** | **Effect size (95% CI)†** | ***p* *value*‡** |
| **Trial contribution:** | | | | | | | | |
| ARPEGGIO | 159 | 159 | | | 99 | 219 | | |
| **Demographics:** | | | | | | | | |
| Age (years) | 45.09 (7.68) | 47.90 (5.56) | -2.81 (-4.29, -1.33) | <0.001 | 44.80 (8.26) | 47.26 (5.95) | -2.46 (-4.29, -0.64) | 0.009 |
| Sex (% male) | 53.46 | 54.72 | 0.95 (0.60, 1.51) | 0.910 | 55.56 | 53.42 | 1.09 (0.66, 1.81) | 0.808 |
| Height (cm) | 172.12 (9.12) | 171.36 (9.95) | 0.76 (-1.35, 2.87) | 0.479 | 173.23 (9.49) | 171.07 (9.51) | 2.15 (-0.11, 4.42) | 0.064 |
| Weight (kg) | 74.71 (17.78) | 74.10 (13.47) | 0.61 (-2.88, 4.09) | 0.733 | 76.00 (18.13) | 73.68 (14.53) | 2.31 (-1.77, 6.40) | 0.267 |
| Disease duration (years) | 6.89 (5.18) | 8.76 (6.12) | -1.87 (-3.12, -0.62) | 0.004 | 6.25 (4.65) | 8.54 (6.04) | -2.29 (-3.52, -1.07) | <0.001 |
| **Disability Scores:** | | | | | | | | |
| EDSS | 4.70 (0.94) | 4.26 (0.91) | 0.44 (0.24, 0.64) | <0.001 | 4.74 (0.89) | 4.36 (0.95) | 0.38 (0.17, 0.60) | <0.001 |
| FSS-Bowel and Bladder | 1.40 (0.97) | 1.05 (0.84) | 0.35 (0.15, 0.55) | <0.001 | 1.38 (0.97) | 1.16 (0.89) | 0.23 (0.00, 0.45) | 0.049 |
| FSS-Brainstem | 0.90 (0.90) | 1.09 (0.95) | -0.19 (-0.40, 0.01) | 0.062 | 0.89 (0.85) | 1.05 (0.96) | -0.16 (-0.37, 0.05) | 0.147 |
| FSS-Cerebellar | 2.41 (0.73) | 1.80 (0.87) | 0.61 (0.43, 0.79) | <0.001 | 2.55 (0.69) | 1.90 (0.85) | 0.64 (0.46, 0.82) | <0.001 |
| FSS-Cerebral | 0.92 (0.92) | 0.87 (0.87) | 0.05 (-0.15, 0.25) | 0.619 | 0.93 (0.91) | 0.89 (0.89) | 0.04 (-0.17, 0.26) | 0.694 |
| FSS-Pyramidal | 2.83 (0.67) | 2.95 (0.51) | -0.12 (-0.25, 0.01) | 0.075 | 2.82 (0.67) | 2.92 (0.56) | -0.10 (-0.26, 0.05) | 0.181 |
| FSS-Sensory | 1.76 (1.03) | 1.71 (1.02) | 0.05 (-0.18, 0.28) | 0.664 | 1.71 (1.08) | 1.75 (1.00) | -0.04 (-0.29, 0.21) | 0.746 |
| FSS-Visual | 1.39 (1.40) | 0.35 (0.69) | 1.04 (0.80, 1.29) | <0.001 | 1.63 (1.53) | 0.53 (0.86) | 1.10 (0.78, 1.43) | <0.001 |
| Mean T25FW (sec) | 10.24 (9.75) | 9.04 (6.57) | 1.21 (-0.63, 3.04) | 0.198 | 10.34 (10.15) | 9.32 (7.35) | 1.03 (-1.22, 3.27) | 0.370 |
| Mean 9HPT dominant (sec) | 29.95 (13.32) | 26.90 (10.93) | 3.04 (0.35, 5.73) | 0.027 | 31.16 (14.55) | 27.19 (10.88) | 3.98 (0.75, 7.21) | 0.017 |
| Mean 9HPT non-dominant (sec) | 33.85 (20.63) | 27.04 (7.60) | 6.81 (3.37, 10.25) | <0.001 | 36.71 (24.30) | 27.61 (8.65) | 9.10 (4.12, 14.08) | <0.001 |
| **MRI metrics:** | | | | | | | | |
| Gad count | 0.58 (1.80) | 0.11 (0.51) | 0.47 (0.18, 0.76) | 0.002 | 0.74 (2.21) | 0.17 (0.56) | 0.56 (0.12, 1.01) | 0.014 |
| T2 Lesion Volume (mL) | 7.77 (10.89) | 4.03 (5.80) | 3.73 (1.81, 5.66) | <0.001 | 8.35 (11.85) | 4.79 (6.94) | 3.55 (1.02, 6.09) | 0.007 |
| Normalized brain volume (L) | 1.44 (0.10) | 1.47 (0.10) | -0.03 (-0.05, -0.01) | 0.012 | 1.44 (0.10) | 1.47 (0.10) | -0.02 (-0.05, 0.00) | 0.063 |

Values in brackets are standard deviations, unless otherwise specified.

*Percentile threshold for defining responders. The 50th percentile defines responders as the top 50% who are predicted to be most responsive, while the 70th percentile defines them as the top 30%. The non-responders are those who fall below the percentile threshold.

†Effect size is the average difference between responders and non-responders for all covariates except for "sex" which is an odd's ratio (OR).

‡*p* values for continuous and ordinal variables are calculated using a two-sided Welch's t-test due to unequal variances/sample sizes. *p* value for the categorical variable "sex" is calculated using a two-sided Fisher's exact test due to unequal and relatively small sample sizes.

EDSS = Expanded Disability Status Scale; FSS = Functional Systems Score; T25FW = timed 25-foot walk; 9HPT = 9-hole peg test; Gad = Gadolinium-enhancing lesion.
